# Physical activity, sedentary behavior, and sleep in everyday life after childhood cancer – a report from the Swiss Childhood Cancer Survivor Study (SCCSS)-Activity

**DOI:** 10.64898/2026.09.17.26363324

**Authors:** Carina Nigg, Maša Žarković, Dominik Mohr, Christina Schindera, Claudia E Kuehni

**Author notes:** Shared first authors (Carina Nigg, Maša Žarković). Corresponding author: Prof. Dr. med. Claudia Kuehni, Mittelstrasse 43, 3012 Bern, Switzerland.

## Abstract

**Background:** Childhood cancer survivors (CCS) are at elevated risk of chronic health conditions. Sufficient physical activity and sleep, and limited sedentary behavior mitigate those risks, but device-based assessments of these behaviors among CCS are scarce. We described physical activity, sedentary behavior, and sleep among CCS using thigh-worn accelerometers and examined associated factors.

**Methods:** We included participants of the 2024-2025 Swiss Childhood Cancer Survivor Study questionnaire wave who consented to wear an activPAL4^®^ accelerometer continuously for eight days. Using ActiPASS, we derived physical activity types and intensities, sedentary behavior, and sleep. We stratified analyses by age (children: 5-17 years; adults: ≥18 years).

**Results:** 108 CCS (68 children, 40 adults) pariticipated. Children accumulated a median of 85 minutes/day of moderate-to-vigorous physical activity [IQR 57-107] and 578 minutes/day sedentary behavior [516-625]; adults 78 [61-100] and 603 minutes/day [522-650]. Both groups spent 4-5 hours/day in sedentary bouts ≥30 minutes. Children slept 466 minutes/night [418-499], and adults 435 [407-488]. Only 9% of children and 20% of adults met recommendations for physical activity, sedentary behavior, and sleep simultaneously. Radiotherapy was associated with lower physical activity among children and higher sedentary time among children and adults. Older children achieved less physical activity and sleep, and had more sedentary time.

**Conclusion:** CCS engage in extensive sedentary behavior, and especially in long uninterrupted sedentary bouts. Many sleep less than recommended. Healthcare practitioners should prioritize promoting physical activity among adolescent survivors and particularly those treated with radiotherapy, and support all survivors in improving sleep duration and reducing sedentary time.

## Introduction

Medical advances in childhood cancer treatment have increased 5-year survival to nearly 90%, but these gains come with substantial long-term morbidity.^1^ By age 50, childhood cancer survivors (CCS) develop on average five serious chronic health conditions, compared to two in the general population, including cardiovascular, pulmonary, and neoplastic events.^2^ A healthy lifestyle, including sufficient physical activity and sleep and limited sedentary behavior, reduces the risk of subsequent neoplasms, cardiovascular, and pulmonary events.^3–6^ However, cancer treatments and their late effects may themselves affect physical activity, sedentary behavior, and sleep.^7,8^ To date, research has mostly focused on self-reported physical activity^9,10^, which is prone to recall and social desirability bias.^11^ Sedentary behavior has been less studied among CCS and predominantly operationalized as self-reported leisure screen-time,^12,13^ neglecting important sedentary periods (e.g., at school or work).^3^ Although sleep disturbances are common in CCS,^14^ studies on sleep duration are scarce and mainly based on self-report.^15,16^

In the general population, the last two decades have seen a shift toward assessing physical activity, sedentary behavior, and sleep in everyday life using wearable technologies such as accelerometers.^17,18^ Accelerometers allow collecting intensive longitudinal data on these behaviors over longer periods in real time in daily life, eliminating recall bias and improving accuracy.^19,20^ Compared with questionnaires, accelerometer-assessed physical activity shows stronger associations with health indicators.^21^ Yet accelerometer studies among CCS remain scarce. The Swiss Childhood Cancer Survivor Study (SCCSS)-Activity addresses this research gap by using thigh-worn accelerometers worn 24 hours per day for one week. We distinguish accelerometer-assessed physical activity types and intensities, sedentary behavior, and sleep, and evaluate which factors are associated with these outcomes.

## Methods

### Study design and study population

The SCCSS-Activity is nested within the national Swiss Childhood Cancer Survivor Study (SCCSS), a population-based questionnaire study of long-term health among all childhood cancer survivors who survived ≥5 years post-diagnosis and who are registered in the Swiss Childhood Cancer Registry (ChCR). The ChCR includes all individuals diagnosed before age 20 with leukemia, lymphoma, CNS tumors, malignant solid tumors, or Langerhans cell histiocytosis in Switzerland since 1976.^22^ Detailed methods of the SCCSS^23^ and SCCSS-Activity^24^ are available elsewhere. In SCCSS-Activity, we included survivors of any age who participated in the 2024-2025 SCCSS questionnaire mailing and indicated interest in an accelerometer study. The study is registered with Clinicaltrials.gov (NCT03297034). Ethical approval for SCCSS and SCCSS-Activity was granted by the ethics committee of the canton of Bern (KEK-BE: 166/2014, 2021-01462, 2024-02240).

### Study procedures SCCSS-Activity

In the SCCSS questionnaire, we asked participants if they would be interested in wearing an accelerometer for one week to objectively assess physical activity (yes/no). After receiving detailed information and providing informed consent, participants received an accelerometer package by post. The package contained the accelerometer, adhesives and replacements, illustrated mounting/dismounting instructions, a wear time protocol, a prepaid return envelope, and a return checklist. We instructed participants to attach the device to the front of the right thigh midway between knee and hip with PALpatch® adhesives. We pre-packed each accelerometer in a nitrile sleeve and sealed it with PALpatch® materials. Participants applied an Opsite Flexifix® adhesive over the device to ensure waterproofing and prevent detachment (e.g., when changing clothes). We instructed participants to mount the accelerometer immediately upon receipt, to wear it continuously for eight days, to remove it on the ninth day in the morning, and to return it by mail. To support correct and timely mounting, we contacted participants the day after they received the package to clarify any uncertainties. Participants who wore the device for at least 20 hours on seven days received a 20 CHF gift card.

### Accelerometer and data processing

We used activPAL4® devices (PAL Technologies, Glasgow, Scotland) for behavioral assessment. The activPAL® is a small and light-weight research-grade device (26x43x5 mm; 9g) with a battery capacity of up to 14 days and 64 MB memory, sampling acceleration on three axes with ±4 g. The activPAL is a valid device to capture posture and activity type in children, adolescents, and adults.^25,26^

We used the PAL Software Suite (v9.1.2.2) to download and export raw data as compressed csv-files for further processing. We processed raw accelerometer data using the validated open-source software ActiPASS (version 2025.10.1). ActiPASS classifies behaviors in 2-second windows with 50% overlap, yielding a one-second epoch resolution, and applies algorithms for detecting non-wear periods, sleep, posture, and activity intensity (cadence-based intensity).^27–31^ ActiPASS has shown high accuracy for wake-time movement behaviors (over 90%) and sleep detection (84%).^28–30,32–34^ Because cadence thresholds are based on adult cadence, we implemented age-specific cadence thresholds for survivors <18 years based on heuristic recommendations.^35^ We visually inspected cases flagged by ActiPASS criteria (e.g., very little walking) for plausibility. If data were implausible, we removed that day from analysis but retained remaining valid days. We further processed daily ActiPASS output using R and RStudio to identify valid datasets and compute weekly averages of our outcomes. We defined a valid dataset as ≥ 20 hours wear time on at least four weekdays and one weekend day.^36,37^

### Physical activity, sedentary behavior, and sleep

#### Physical activity types

We derived time spent in walking, running, stair walking (including terrain walking), cycling, moving (standing posture with intermittent steps), other activity (not classifiable into the above, e.g., gym exercises), and a composite “movement” variable (sum of all activity types). We also calculated mean daily steps.

#### Physical activity intensities

Light physical activity defines activities requiring 1.5-3 metabolic equivalents (METs).^38,39^ We computed light physical activity with standing (ambulatory movement without purposeful walking with a cadence <100/min; slow walking as defined by age-specific thresholds) and without standing. Moderate-to-vigorous physical activity (MVPA) defines activities requiring ≥ 3 METs (fast walking, running, cycling, stair walking, ambulatory movement without purposeful walking with a cadence ≥ 100/min).

#### Sedentary behavior and sedentary bouts

Sedentary behavior is defined as sitting or lying while awake and requiring < 1.5 METs.^38^ Because uninterrupted sedentary bouts ≥ 30 minutes are particularly detrimental to health,^40^ we calculated frequencies of and time spent in sedentary bouts of ≥ 30 bouts.

#### Sleep

We defined sleep as total sleep time within each time-in-bed period, which was based on long and continuous lying periods.

#### Adherence to guidelines and recommendations

We defined physical activity guideline adherence based upon the World Health Organization (WHO): ≥ 60 minutes MVPA/day for children, and ≥150 minutes MVPA/week for adults.^4^ Sleep guidelines adherence was defined based upon the National Sleep Science Foundation: 9-11 hours/night for 6-13 years year-olds, 8-10 hours/night for 14-17 year-olds, and 7-9 hours/night for adults.^41^ In the absence of a consensus on maximal sedentary time, we considered adherence to sedentary behavior recommendations as < 9 hours/day sedentary for adults based on evidence of sedentary behavior and all-cause mortality.^42^ For children, recommendations exist only for recreational screen time and avoiding prolonged sitting.^43^ We considered < 8 hours/day of sedentary time as adhering to sedentary behavior recommendations based on adult sedentary recommendations and the assumption that children should engage in 4–5 hours more MVPA per week than adults.^4,44^

### Explanatory variables

#### Sociodemographic characteristics

We assessed sociodemographic characteristics via questionnaires: Swiss language region, parental and participant nationality and country of birth, and parental education. We categorized language region as German versus French/Italian. Definitions for migration background and parental education followed the Swiss Federal Statistical Office.^45,46^ If both parents were born Swiss or if the participant and at least one parent were born Swiss, we defined this as no migration background.^45^ For educational background, we used the highest educational level attained by either parent and categorized it as primary (compulsory schooling only), secondary (high schooling or vocational training), or tertiary (advanced vocational education, university, or technical college).^46^ We coded place of residence as city, town, or rural according to the Federal Statistical Office urban–rural typology.^47^

#### Cancer-related characteristics

We received the following cancer-related characteristics from the ChCR: sex, date of birth, age at diagnosis, and diagnosis according to the International Classification of Childhood Cancer, third edition (ICCC-3).^48^ Treatment information included radiotherapy, chemotherapy, surgery, HSCT, and relapse (all no/yes).

#### Exercise and active travel

In the SCCSS questionnaire, we asked participants whether they engage in exercise and whether any problems hinder them from exercising (yes/no). We asked children <16 years how they usually commute to school and those ≥16 years how they usually commute in everyday life (e.g., to school or work). Response options were walking; (e-)bicycle, skateboard, or scooter; public transport; and car, moped, or e-scooter. Multiple modes could be selected. We categorized participants reporting walking or (e-)bicycle, skateboard, or scooter as using active transportation.

### Statistical analysis

We compared SCCSS-Activity responders and non-responders (including those without valid accelerometer data) using descriptive statistics, chi-square tests, and t-tests. All subsequent analyses were stratified by age group (children: 5–17 years; adults: ≥18 years). We used descriptive statistics to analyze time spent in physical activity types and intensities, standing, sedentary behavior and sedentary bouts, and sleep. To facilitate comparability, we applied post-stratified weighting for children and adolescents by age and sex using published results from the German Motorik-Modul (MoMo) Study, which used the same device, protocol, and data processing procedures as SCCSS-Activity.^49^ We also computed the proportion of time CCS spent in sedentary behavior, light physical activity, and MVPA relative to time awake and to accelerometer wear time to allow comparison with studies using wake-time-only protocols.^3^ We calculated the number and proportion of children and adults adhering to physical activity, sedentary behavior, and sleep recommendations. To explore associated factors, we selected the following outcomes as key activity indicators: movement (all physical activity types irrespective of intensity), steps, light physical activity, MVPA, sedentary behavior, and sleep.^4,50^ We included age, sex, parental education, and migration, *a priori* as important physical activity confounders.^51,52^ We added treatment-related variables (radiotherapy, chemotherapy, HSCT, surgery) as primary exposures of interest. Due to the limited sample size and the exploratory nature of the analyses, we focused on estimation rather than formal hypothesis testing. We report both unstandardized (B) and standardized (β) regression coefficients to compare the relative strength of predictors measured on different scales. For result interpretation, we considered the magnitude and precision of estimated effects instead of solely relying on conventional p-value thresholds (p<0.05).

## Results

In total, 369 CCS responded to the SCCSS questionnaire. Of those, 157 indicated an interest, 117 participated, and 108 (29%) provided valid accelerometer data (*Supplementary Figure 1*), including 68 children and 40 adults. Median age at study was 14 years (interquartile range [IQR] 11-21), at diagnosis 6 years [IQR 3-13], and time since diagnosis 8 years [IQR 7-9] (*Table 1*). Half were female, and 69% had parents with tertiary education. Leukemias were the most frequent diagnosis among pediatric survivors (43%) and lymphomas among adults (28%). Overall, 72% had received chemotherapy, 27% surgery, 24% radiotherapy, and 8% HSCT. Participants were largely similar to non-participants but more often reported using active commuting (*Supplementary Table 1*). Survivors wore the accelerometer for a median of 7 days [IQR 7-8], 24 hours/day (*Table 2*).

**Table 1.** Characteristics of SCCSS-Activity study participants.

|  | **Children**<br>**(5-17 years)**<br>**N = 68** | **Adults**<br>**(≥18 years)**<br>**N = 40** | **Overall**<br>**N = 108** |
| --- | --- | --- | --- |
| **Socio-demographic characteristics** | **n (%)** | **n (%)** | **n (%)** |
| *Age at study [median, IQR]* | 12 [10-14] | 24 [21-27] | 14 [11-21] |
| *Female* | 27 (40%) | 27 (68%) | 54 (50%) |
| *Language* |  |  |  |
| German | 49 (72%) | 29 (72%) | 78 (72%) |
| French/Italian | 19 (28%) | 11 (28%) | 30 (28%) |
| *Migration background* | 11 (16%) | 7 (18%) |  |
| *Parental education* |  |  |  |
| Primary education | 0 (0%) | 1 (3%) | 1 (1%) |
| Secondary education | 10 (15%) | 22 (55%) | 32 (30%) |
| Tertiary education | 58 (85%) | 17 (43%) | 75 (69%) |
| *Living location* |  |  |  |
| Rural | 9 (13%) | 6 (15%) | 15 (14%) |
| Town | 24 (35%) | 10 (25%) | 34 (31%) |
| City | 35 (52%) | 24 (60%) | 59 (55%) |
| **Questionnaire-reported physical activity** |  |  |  |
| Engages in exercise | 63 (93%) | 29 (74%) | 92 (87%) |
| Physical problem hindering exercise | 5 (8%) | 9 (23%) | 14 (13%) |
| Active travel | 57 (84%) | 30 (75%) | 87 (81%) |
| **Cancer-related characteristics** |  |  |  |
| *ICCC3 main group* |  |  |  |
| Leukemias | 29 (43%) | 8 (20%) | 37 (34%) |
| Lymphomas | 3 (4%) | 11 (28%) | 14 (13%) |
| CNS tumors | 15 (22%) | 9 (23%) | 24 (22%) |
| Other | 21 (31%) | 12 (30%) | 33 (31%) |
| *Age at diagnosis, median [IQR]* | 4 [2-5] | 15 [13-19] | 6 [3-13] |
| *Time since diagnosis, median [IQR]* | 8 [7-9] | 8 [7-9] | 8 [7-9] |
| *Radiotherapy* | 15 (22%) | 11 (28%) | 26 (24%) |
| *Chemotherapy* | 55 (81%) | 23 (58%) | 78 (72%) |
| *Surgery* | 19 (28%) | 10 (25%) | 29 (27%) |
| *HSCT* | 7 (10%) | 1 (3%) | 8 (8%) |
| *Relapse* | 0 (0%) | 0 (0%) | 0 (0%) |

ICCC3 = International Classification of Childhood Cancer, 3<sup>rd</sup> edition, CNS = Central nervous system, HSCT = Hematopoietic stem cell transplantation, p-value is based upon chi-square and t-test test

**Table 2.**
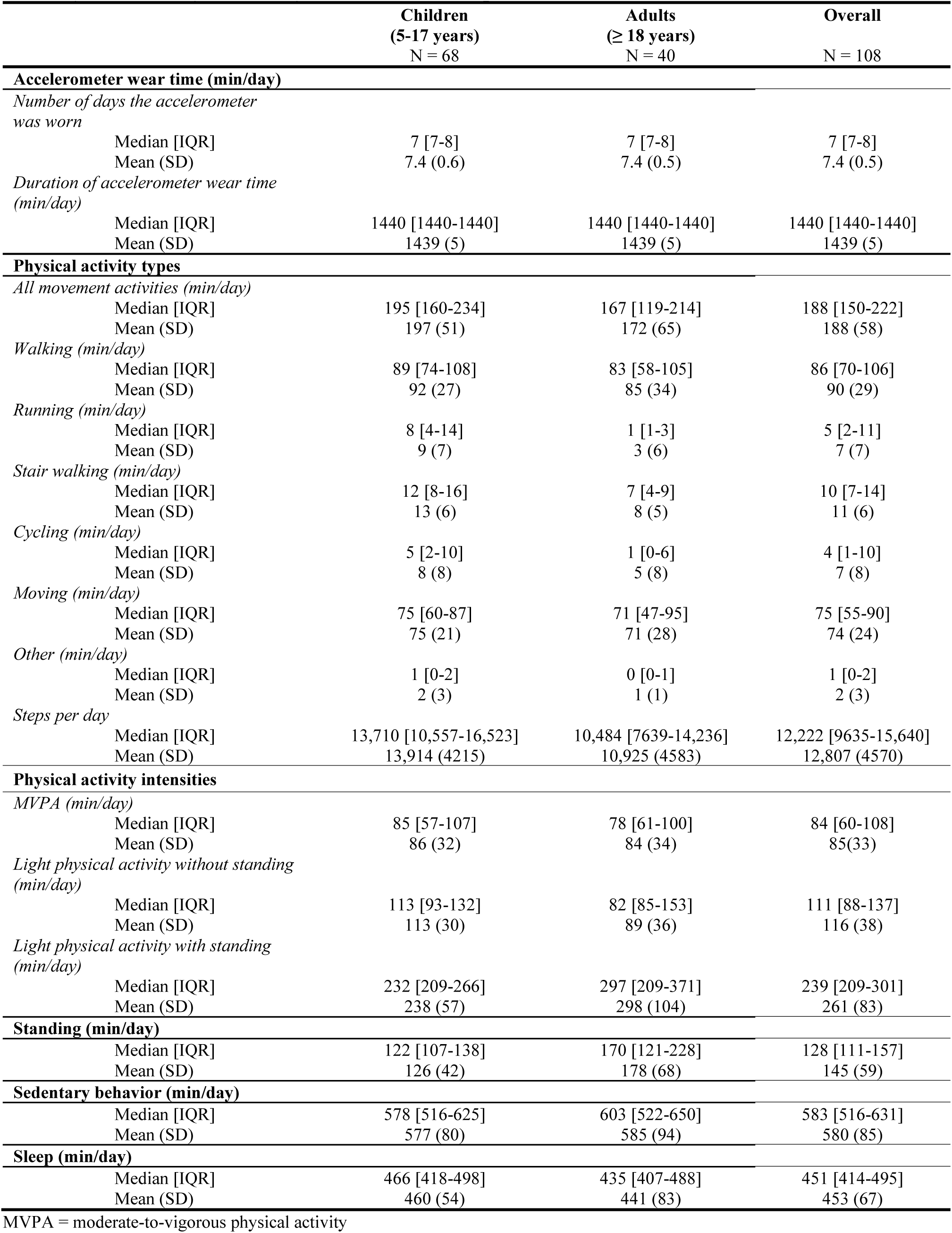
Physical activity, sedentary behavior, and sleep of childhood cancer survivors.

|  | Children<br>(5-17 years)<br>N = 68 | Adults<br>(≥ 18 years)<br>N = 40 | Overall<br>N = 108 |
| --- | --- | --- | --- |
| **Accelerometer wear time (min/day)** |  |  |  |
| *Number of days the accelerometer was worn* |  |  |  |
| Median [IQR] | 7 [7-8] | 7 [7-8] | 7 [7-8] |
| Mean (SD) | 7.4 (0.6) | 7.4 (0.5) | 7.4 (0.5) |
| *Duration of accelerometer wear time (min/day)* |  |  |  |
| Median [IQR] | 1440 [1440-1440] | 1440 [1440-1440] | 1440 [1440-1440] |
| Mean (SD) | 1439 (5) | 1439 (5) | 1439 (5) |
| **Physical activity types** |  |  |  |
| *All movement activities (min/day)* |  |  |  |
| Median [IQR] | 195 [160-234] | 167 [119-214] | 188 [150-222] |
| Mean (SD) | 197 (51) | 172 (65) | 188 (58) |
| *Walking (min/day)* |  |  |  |
| Median [IQR] | 89 [74-108] | 83 [58-105] | 86 [70-106] |
| Mean (SD) | 92 (27) | 85 (34) | 90 (29) |
| *Running (min/day)* |  |  |  |
| Median [IQR] | 8 [4-14] | 1 [1-3] | 5 [2-11] |
| Mean (SD) | 9 (7) | 3 (6) | 7 (7) |
| *Stair walking (min/day)* |  |  |  |
| Median [IQR] | 12 [8-16] | 7 [4-9] | 10 [7-14] |
| Mean (SD) | 13 (6) | 8 (5) | 11 (6) |
| *Cycling (min/day)* |  |  |  |
| Median [IQR] | 5 [2-10] | 1 [0-6] | 4 [1-10] |
| Mean (SD) | 8 (8) | 5 (8) | 7 (8) |
| *Moving (min/day)* |  |  |  |
| Median [IQR] | 75 [60-87] | 71 [47-95] | 75 [55-90] |
| Mean (SD) | 75 (21) | 71 (28) | 74 (24) |
| *Other (min/day)* |  |  |  |
| Median [IQR] | 1 [0-2] | 0 [0-1] | 1 [0-2] |
| Mean (SD) | 2 (3) | 1 (1) | 2 (3) |
| *Steps per day* |  |  |  |
| Median [IQR] | 13,710 [10,557-16,523] | 10,484 [7639-14,236] | 12,222 [9635-15,640] |
| Mean (SD) | 13,914 (4215) | 10,925 (4583) | 12,807 (4570) |
| **Physical activity intensities** |  |  |  |
| *MVPA (min/day)* |  |  |  |
| Median [IQR] | 85 [57-107] | 78 [61-100] | 84 [60-108] |
| Mean (SD) | 86 (32) | 84 (34) | 85(33) |
| *Light physical activity without standing (min/day)* |  |  |  |
| Median [IQR] | 113 [93-132] | 82 [85-153] | 111 [88-137] |
| Mean (SD) | 113 (30) | 89 (36) | 116 (38) |
| *Light physical activity with standing (min/day)* |  |  |  |
| Median [IQR] | 232 [209-266] | 297 [209-371] | 239 [209-301] |
| Mean (SD) | 238 (57) | 298 (104) | 261 (83) |
| **Standing (min/day)** |  |  |  |
| Median [IQR] | 122 [107-138] | 170 [121-228] | 128 [111-157] |
| Mean (SD) | 126 (42) | 178 (68) | 145 (59) |
| **Sedentary behavior (min/day)** |  |  |  |
| Median [IQR] | 578 [516-625] | 603 [522-650] | 583 [516-631] |
| Mean (SD) | 577 (80) | 585 (94) | 580 (85) |
| **Sleep (min/day)** |  |  |  |
| Median [IQR] | 466 [418-498] | 435 [407-488] | 451 [414-495] |
| Mean (SD) | 460 (54) | 441 (83) | 453 (67) |

MVPA = moderate-to-vigorous physical activity

### Time spent in physical activity, sedentary behavior, and sleep

Children spent a median of 195 min/day (3.3 h/day) on any physical activity, and adults 172 min/day (2.8 h/day) (*Table 2*). Walking contributed most to physical activity (children: 89 minutes/day [IQR 74-108); adults: 83 minutes/day [IQR 58-105]). Children accumulated 85 minutes/day of MVPA [IQR 57-107], equivalent to 10% [IQR 6-12] of awake time, and adults 78 minutes/day [IQR 61-100], equivalent to 8% [IQR 7-11] of awake time (*Supplementary Table 2*). Children spent a median of 578 min/day (9.6 h/day) sedentary (64% [IQR 58-69] of awake time) and slept 466 min/day (7.8 h/day). Adults spent 603 min/day (10 h/day) sedentary (60% [IQR 55-73] or awake time) and slept 583 min/day (7.3 h/day). Weighted descriptive results for comparison with the MoMo-Study are provided in *Supplementary Table 3*. Children accumulated 226 minutes/day [IQR 159-320] (4 hours/day) and adults 319 minutes/day [IQR 230-416] (5 hours/day) in sedentary bouts ≥30 minutes (*Supplementary Table 4*).

### Guidelines

Overall, 72% (95%CI 60-82) of children met the WHO’s physical activity recommendations, as did 98% (95%CI 83-100) of adults (*Figure 1, Supplementary Table 5*). Forty-three percent (95%CI 31-55) of children and 65% (95%CI 48-79) of adults adhered to sleep recommendations. Sedentary behavior adherence was lowest: 28% of adults (95% CI 16–44) spent <9 hours/day sedentary, and 10% of children (95% CI 5–20) spent <8 hours/day sedentary. Only 9% of children (95%CI 4-19) and 20% of adult CCS (95%CI 10-36) adhered to physical activity, sedentary behavior, and sleep guidelines simultaneously.

**Figure 1.**
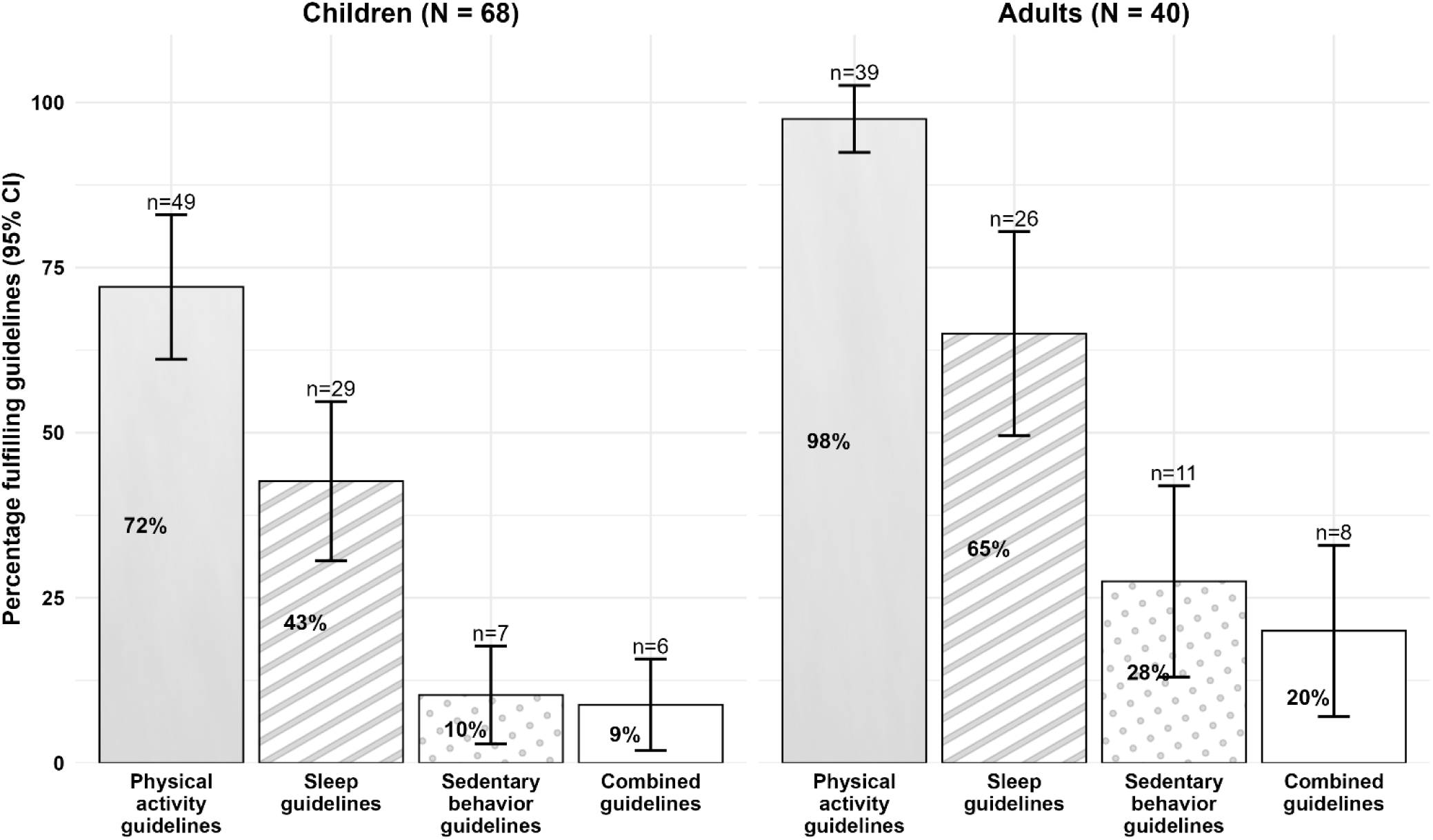
Adherence to physical activity, sedentary behavior, and sleep recommendations, and all guidelines combined among pediatric and adult childhood cancer survivors. Physical activity adherence was defined according to WHO guidelines [4]. Sleep adherence was defined according to National Sleep Foundation recommendations [41]. Sedentary behavior adherence was defined as <9 h/day in adults based on evidence relating sedentary time to all-cause mortality [42] and as <8 h/day in children based on pediatric sedentary behavior recommendations and study-specific assumptions [43, 44].

### Factors associated with physical activity types and intensities

Among children, radiotherapy was consistently associated with less physical activity: -1990 steps/day (95%CI -4442; 463, β = -0.47), -35.59 minutes/day of any movement activities (95%CI -64.06; -7.12, β = -0.70), and -25.80 minutes/day of light physical activity (95%CI - 44.08; -7.69, β = -0.87) (*Table 3*). Associations with MVPA were less clear (B = -9.52, 95%CI -25.21; 6.17, β = -0.30). For each year of age, children accumulated -557 steps/day (95%CI -900; -215, β = -0.37) and spent -6.74 minutes/day in any movement activity (95%CI -10.71; 2.76, β = -0.37) and -6.99 minutes/day in MVPA (95%CI -9.18; -4.80, β = -0.61).

**Table 3.** Factors associated with physical activity type and intensity in childhood cancer survivors based on multivariable linear regression models.

|  | Children (5-17 years) |  |  |  | Adults (≥ 18 years) |  |  |  |
| --- | --- | --- | --- | --- | --- | --- | --- | --- |
|  | N = 68 |  |  |  | N = 40 |  |  |  |
| | *B* | 95%CI | $\beta$ | *p* | *B* | 95%CI | $\beta$ | *p* |
| **Steps (per day)** |  |  |  |  |  |  |  |  |
| *Intercept* | 17830 | 12011;23649 | -0.62 | **<0.001** | 8630 | -6099;23359 | 0.02 | 0.241 |
| Age at study | -557 | -900;-215 | -0.37 | **0.002** | 100 | -487;687 | 0.08 | 0.731 |
| Male<sup>a</sup> | 1788 | -125;3701 | 0.42 | 0.066 | -351 | -3956;3253 | -0.08 | 0.844 |
| Tertiary parental education<sup>c</sup> | 1809 | -898;4517 | 0.43 | 0.186 | -371 | -3830;3087 | -0.08 | 0.828 |
| Migration background<sup>d</sup> | -555 | -3164;2054 | -0.13 | 0.672 | -889 | -5573;3794 | -0.19 | 0.701 |
| Radiotherapy<sup>e</sup> | -1990 | -4442;463 | -0.47 | 0.110 | -655 | -5249;3940 | -0.14 | 0.773 |
| Chemotherapy<sup>f</sup> | 970 | -1643;3582 | 0.23 | 0.461 | -67 | -3821;3688 | -0.01 | 0.971 |
| Surgery<sup>g</sup> | -372 | -2430;1686 | -0.09 | 0.719 | 811 | -3057;4680 | 0.18 | 0.672 |
| HSCT<sup>h</sup> | -310 | -3631;3011 | -0.07 | 0.853 |  |  |  |  |
| **Any movement activity (minutes/day)** |  |  |  |  |  |  |  |  |
| *Intercept* | 252.18 | 184.65;319.72 | -0.47 | **<0.001** | 96.93 | -109.31;303.17 | -0.15 | 0.346 |
| Age at study | -6.74 | -10.71;-2.76 | -0.37 | **0.001** | 2.72 | -5.50;10.94 | 0.15 | 0.505 |
| Male<sup>a</sup> | 18.44 | -3.76;40.65 | 0.36 | 0.102 | 8.13 | -42.35;58.60 | 0.13 | 0.745 |
| Tertiary parental education<sup>c</sup> | 21.60 | -9.83;53.02 | 0.42 | 0.174 | 6.33 | -42.11;54.76 | 0.10 | 0.792 |
| Migration background<sup>d</sup> | -22.31 | -52.58;7.97 | -0.44 | 0.146 | -11.32 | -76.90;54.26 | -0.17 | 0.727 |
| Radiotherapy<sup>e</sup> | -35.59 | -64.06;-7.12 | -0.70 | **0.015** | -31.12 | -95.45;33.22 | -0.48 | 0.332 |
| Chemotherapy<sup>f</sup> | 8.59 | -21.73;38.90 | 0.17 | 0.573 | 11.66 | -40.91;64.23 | 0.18 | 0.655 |
| Surgery<sup>g</sup> | -3.95 | -27.83;19.94 | -0.08 | 0.742 | 12.22 | -41.95;66.39 | 0.19 | 0.649 |
| HSCT<sup>h</sup> | 13.47 | -25.07;52.01 | 0.26 | 0.487 |  |  |  |  |
| **Light physical activity (minutes/day)** |  |  |  |  |  |  |  |  |
| *Intercept* | 105.61 | 62.44;148.78 | -0.36 | **<0.001** | 19.90 | -91.98;131.78 | -0.32 | 0.720 |
| Age at study | -0.28 | -2.83;2.26 | -0.03 | 0.824 | 2.39 | -2.07;6.85 | 0.23 | 0.283 |
| Male<sup>a</sup> | 11.76 | -2.43;25.95 | 0.40 | 0.103 | 14.52 | -12.85;41.90 | 0.40 | 0.288 |
| Tertiary parental education<sup>c</sup> | 12.77 | -7.31;32.86 | 0.43 | 0.208 | 7.43 | -18.84;33.70 | 0.21 | 0.569 |
| Migration background<sup>d</sup> | -15.71 | -35.07;3.64 | -0.53 | 0.110 | 4.11 | -31.46;39.68 | 0.11 | 0.815 |
| Radiotherapy<sup>e</sup> | -25.89 | -44.08;-7.69 | -0.87 | **0.006** | -24.29 | -59.19;10.61 | -0.67 | 0.166 |
| Chemotherapy<sup>f</sup> | -1.83 | -21.21;17.55 | -0.06 | 0.851 | 13.24 | -15.28;41.76 | 0.37 | 0.351 |
| Surgery<sup>g</sup> | 2.35 | -12.92;17.62 | 0.08 | 0.759 | 3.06 | -26.32;32.45 | 0.08 | 0.833 |
| HSCT<sup>h</sup> | 9.64 | -15.00;34.28 | 0.33 | 0.437 |  |  |  |  |
| **Moderate-to-vigorous physical activity (minutes/day)** |  |  |  |  |  |  |  |  |
| *Intercept* | 157.05 | 119.82;194.28 | -0.36 | **<0.001** | 77.35 | -30.59;185.29 | 0.05 | 0.154 |
| Age at study | -6.99 | -9.18;-4.80 | -0.61 | **<0.001** | 0.35 | -3.96;4.65 | 0.03 | 0.871 |
| Male<sup>a</sup> | 7.97 | -4.27;20.20 | 0.25 | 0.198 | -6.46 | -32.88;19.95 | -0.19 | 0.622 |
| Tertiary parental education<sup>c</sup> | 6.28 | -11.04;23.60 | 0.20 | 0.471 | -1.89 | -27.24;23.45 | -0.06 | 0.880 |
| Migration background<sup>d</sup> | -5.43 | -22.12;11.26 | -0.17 | 0.517 | -16.13 | -50.45;18.19 | -0.47 | 0.346 |
| Radiotherapy<sup>e</sup> | -9.52 | -25.21;6.17 | -0.30 | 0.230 | -7.36 | -41.04;26.31 | -0.21 | 0.659 |
| Chemotherapy<sup>f</sup> | 9.81 | -6.90;26.52 | 0.31 | 0.245 | -1.42 | -28.93;26.09 | -0.04 | 0.917 |
| Surgery<sup>g</sup> | -7.52 | -20.69;5.64 | -0.24 | 0.258 | 9.94 | -18.41;38.29 | 0.29 | 0.480 |
| HSCT<sup>h</sup> | 6.14 | -15.11;27.39 | 0.19 | 0.565 |  |  |  |  |

<sup>a</sup>reference: female, <sup>b</sup>reference: German language region, <sup>c</sup>reference: secondary education, <sup>d</sup>reference: no migration background, <sup>e</sup>reference: no radiotherapy; <sup>f</sup>reference: no chemotherapy, <sup>g</sup>reference: no surgery, <sup>h</sup>reference: no HSCT; HSCT = hematopoietic stem cell transplantation, B = unstandardized beta coefficient, $\beta$ = standardized beta coefficient

Boys accumulated more steps than girls (B = 1788, 95%CI -125; 3701, β = -0.42). Among adults, sociodemographic and cancer-related factors were unrelated physical activity outcomes.

### Factors associated with sedentary behavior and sleep

Children treated with radiotherapy spent 38.26 more minutes/day sedentary (95%CI -3.16; 79.68, β = 0.48), and adults 93.46 minutes/day (95%CI 13.87; 173.05, β = 0.99) (*Table 4*). Adults who had surgery spent less time sedentary (B = -67.55, 95%CI -134.56; -0.53, β = -0.72). Associations with age differed by group: per year older, children spent 15.09 minutes/day more sedentary (95% CI 9.30; 20.88, β = 0.52), whereas adults spent 9.07 minutes/day less sedentary (95% CI -19.24; 1.11, β = −0.33). Boys spent more sedentary time than girls (B = 31.85 minutes/day, 95% CI −0.46; 64.16, β = 0.40). Children with tertiary-educated parents spent less sedentary time (B = -55.33 minutes/day, 95% CI -101.05; -9.61, β = -0.69) compared with those with primary/secondary education.

**Table 4.** Factors associated with sedentary behavior and sleep among childhood cancer survivors based on multivariable linear regression models.

| | Children & adolescents (5-17 years) | | | | Adults ($\geq 18$ years) | | | |
| --- | --- | --- | --- | --- | --- | --- | --- | --- |
|  | N = 68 |  |  |  | N = 40 |  |  |  |
| | *B* | *95%CI* | $\beta$ | *p* | *B* | *95%CI* | $\beta$ | *p* |
| Sedentary behavior (minutes/day) |  |  |  |  |  |  |  |  |
| *Intercept* | 446.02 | 347.75;544.28 | 0.58 | **<0.001** | 806.56 | 551.42;1061.71 | 0.05 | **<0.001** |
| Age at study | 15.09 | 9.30;20.88 | 0.52 | **<0.001** | -9.07 | -19.24;1.11 | -0.33 | 0.079 |
| Male<sup>a</sup> | 31.85 | -0.46;64.16 | 0.40 | 0.053 | -24.18 | -86.62;38.26 | -0.26 | 0.436 |
| Tertiary parental education<sup>c</sup> | -55.33 | -101.05;-9.61 | -0.69 | **0.019** | 25.57 | -34.34;85.49 | 0.27 | 0.391 |
| Migration background<sup>d</sup> | 21.09 | -22.97;65.14 | 0.26 | 0.342 | 35.93 | -45.20;117.05 | 0.38 | 0.374 |
| Radiotherapy<sup>e</sup> | 38.26 | -3.16;79.68 | 0.48 | 0.070 | 93.46 | 13.87;173.05 | 0.99 | **0.023** |
| Chemotherapy<sup>f</sup> | -29.20 | -73.31;14.92 | -0.36 | 0.190 | 10.22 | -54.81;75.26 | 0.11 | 0.751 |
| Surgery<sup>g</sup> | -11.02 | -45.78;23.73 | -0.14 | 0.528 | -67.55 | -134.56;-0.53 | -0.72 | **0.048** |
| HSCT<sup>h</sup> | -4.09 | -60.17;52.00 | -0.05 | 0.885 |  |  |  |  |
| Sleep |  |  |  |  |  |  |  |  |
| *Intercept* | 535.71 | 459.35;612.07 | -0.28 | **<0.001** | 472.56 | 222.08;723.03 | -0.23 | **0.001** |
| Age at study | -7.72 | -12.22;-3.22 | -0.39 | **0.001** | -2.13 | -12.12;7.86 | -0.09 | 0.667 |
| Male<sup>a</sup> | -16.67 | -41.77;8.44 | -0.31 | 0.189 | -13.30 | -74.60;48.00 | -0.16 | 0.661 |
| Tertiary parental education<sup>c</sup> | 10.61 | -24.92;46.14 | 0.20 | 0.552 | -31.79 | -90.61;27.03 | -0.38 | 0.279 |
| Migration background<sup>d</sup> | 30.80 | -3.43;65.04 | 0.57 | 0.077 | -17.81 | -97.45;61.84 | -0.21 | 0.652 |
| Radiotherapy<sup>e</sup> | 19.64 | -12.55;51.83 | 0.36 | 0.227 | -29.84 | -107.98;48.30 | -0.36 | 0.442 |
| Chemotherapy<sup>f</sup> | 12.53 | -21.75;46.81 | 0.23 | 0.468 | 10.84 | -53.01;74.69 | 0.13 | 0.732 |
| Surgery<sup>g</sup> | -11.14 | -38.15;15.87 | -0.21 | 0.413 | 62.72 | -3.07;128.51 | 0.75 | 0.061 |
| HSCT<sup>h</sup> | 29.42 | -14.16;73.00 | 0.54 | 0.182 |  |  |  |  |

<sup>a</sup>reference: female, <sup>b</sup>reference: German language region, <sup>c</sup>reference: secondary education, <sup>d</sup>reference: no migration background, <sup>e</sup>reference: no radiotherapy; <sup>f</sup>reference: no chemotherapy, <sup>g</sup>reference: no surgery, <sup>h</sup>reference: no HSCT; HSCT = hematopoietic stem cell transplantation, B = unstandardized beta coefficient

For each one year older, children slept -7.79 minutes/night (95%CI -12.22; -322, β = -0.39). Children with migration background tended to sleep more (B = 30.80 minutes/night, 95%CI - 3.43; 65.04, β = 0.57), as did adults treated with surgery (B = 62.72 minutes/night, 95%CI - 3.07; 128.51, β = 0.75) (*Table 4*).

## Discussion

Survivors of childhood cancer engaged in sufficient physical activity, but spent extensive time sedentary. Most pediatric survivors slept less than recommended. Radiotherapy was associated with lower physical activity among children and higher sedentary behavior in both children and adults. Boys were more sedentary than girls. Older pediatric survivors were less physically active, more sedentary and slept less.

Comparisons with previous CCS studies are challenging because no prior studies have used 24-hour thigh-worn accelerometers in this population. In the multinational PACCS study of 432 CCS aged 6–16 years, survivors accumulated 54 minutes/day of MVPA and 523 minutes/day of sedentary time using hip-worn accelerometers.^53^ In comparison, pediatric survivors in our study accumulated more MVPA (86 minutes/day) but also more sedentary time (577 minutes/day). Adults in our study accumulated 84 minutes/day of MVPA, which is about five times more than the 1223 adult CCS in the SJLIFE cohort assessed with hip-worn accelerometers.^54^ Higher MVPA in our sample likely reflects methodological differences: hip-worn devices are removed during water activities, underestimate cycling, stair climbing, and resistance training, usually require less wear time (≥8 hours), and often use coarser epoch lengths.^53,55^ Contextual factors may also contribute: Switzerland has the highest prevalence of sufficiently active children in Europe,⁵⁶ which may translate to more physical activity among CCS.^56^ Commuting patterns also differ between countries, with walking and cycling being more common in Switzerland, whereas car use predominates in the US.

Compared with the general population, survivors showed similar levels of physical activity, sedentary behavior, and sleep. In the Swiss SOPHYA study of 6-16-year-old children, participants achieved 79 minutes/day of MVPA using hip-worn accelerometers,^57^ comparable to pediatric survivors in our study. Relative to the German Motorik-Modul (MoMo) Study, which used the same 24-hour thigh-worn accelerometry and data processing, pediatric survivors spent slightly less time sedentary and more time in physical activity and sleep.^49^ Among adult CCS, time spent in activity intensities, sleep, and sedentary behavior was similar to results from the 15,253 adults in the international ProPASS consortium using 24-hour thigh-worn accelerometry and the same data processing as this study.^36^

Despite generally comparable behavior patterns with the general population, sedentary behavior and sleep fell short of recommendations. While most survivors met physical activity guidelines, only a minority met all three guidelines simultaneously, mostly due to extensive sedentary time and less than recommended sleep. Survivors spent nearly two-thirds of their awake time sedentary, with about half of this time accumulated in bouts ≥ 30 minutes, which is especially detrimental for health.^40^ This requires further investigation into the types of sedentary behavior and how they relate to survivor’s health.

Sleep duration, although comparable to general population estimates,^36,49,58^ was insufficient for many survivors, warranting further study of sleep disturbances^14,59,60^ and modifiable lifestyle factors^61,62^ to inform sleep interventions.

Radiotherapy showed the strongest and most consistent associations with reduced physical activity in children, contrasting with previous CCS studies that generally found no association,^13,63,64^ although ALL survivors in the Childhood Cancer Survivor Study who received cranial radiation were less likely to be physically active.^65^ Radiotherapy can impair systems essential for physical activity, including musculoskeletal function,^66^ respiratory capacity during physical activity, ^67^ and muscular strength and endurance,^68^ which may lead to less physical activity and more sedentary behavior.

Among pediatric survivors, older age was related to less physical activity, more sedentary behavior, and less sleep, mirroring previous findings,^53,55,58,69^ likely reflecting developmental shifts toward academic demands, social activities, and greater independence, alongside declining access to physical activity-supportive environments.^70,71^ Notably, while age is typically the strongest correlate of physical activity in the general pediatric population,^57,70,72,73^ radiotherapy showed stronger associations than age for most outcomes in our study, suggesting its long-term effects may outweigh typical behavioral correlates. Although boys typically accumulate more physical activity than girls, they were more sedentary in our study, possibly due to higher screen-time engagement.^13^ Among adult survivors, older age was associated with less sedentary time, consistent with prior research,^74^ possibly reflecting increased independence and daily responsibilities that reduce prolonged sitting. Adult survivors who underwent surgery slept longer and were less sedentary; the mechanisms behind this observation require further investigation.

This study advances survivorship research by moving beyond the focus on physical activity by using 24-hour device-based assessment to jointly quantify physical activity, sedentary behavior, and sleep across pediatric, adolescent, and adult survivors. Protocol adherence was high, with 96% providing valid accelerometer data. Limitations include potential self-selection bias, as only participants in the ongoing SCCSS were eligible, and the sample was highly educated. Nevertheless, responders and non-responders were largely comparable. Our adult sample was relatively young, limiting generalizability to older CCS. Limited statistical power restricts the precision of estimates, particularly for weaker associations.

Our findings highlight the need for strategies to reduce sedentary behavior, interrupt prolonged sedentary bouts, and improve sleep duration. In the general population, replacing even 4–12 minutes/day of sedentary time with physical activity can improve cardiometabolic health in children and adults.^36,75^ Evidence on sedentary behavior interventions in CCS is scarce and showed little effect,^76^ and sleep interventions have primarily focused on feasibility rather than efficacy.^77^ Future trials are needed to identify effective strategies for this population. Healthcare practitioners should prioritize promoting physical activity among adolescent survivors, particularly those treated with radiotherapy, and support all survivors in improving sleep duration and reducing sedentary time.

## Statements and Declarations

### Ethics approval

The Ethics Committee of the Canton of Bern (2024-02240) granted ethical approval.

## Acknowledgements

We thank all survivors for participating in our study, the study team of the Childhood Cancer Research Group, the data managers of the Swiss Paediatric Oncology Group, and the team of the Swiss Childhood Cancer Registry.

## Data availability statement

The data that support the information of this manuscript were accessed on secured servers of the Institute of Social and Preventive Medicine at the University of Bern. Individual-level, fully anonymized, sensitive data can only be made available for researchers who fulfil the respective legal requirements. Requests of data from the Childhood Cancer Registry must be directed to the Childhood Cancer Registry of Switzerland (https://www.childhoodcancerregistry.ch). Requests of data from the Swiss Childhood Cancer Survivor Study (SCCSS) and SCCSS-Activity should be communicated to the study lead Claudia E. Kuehni.

---

**Supplementary Figure 1.**
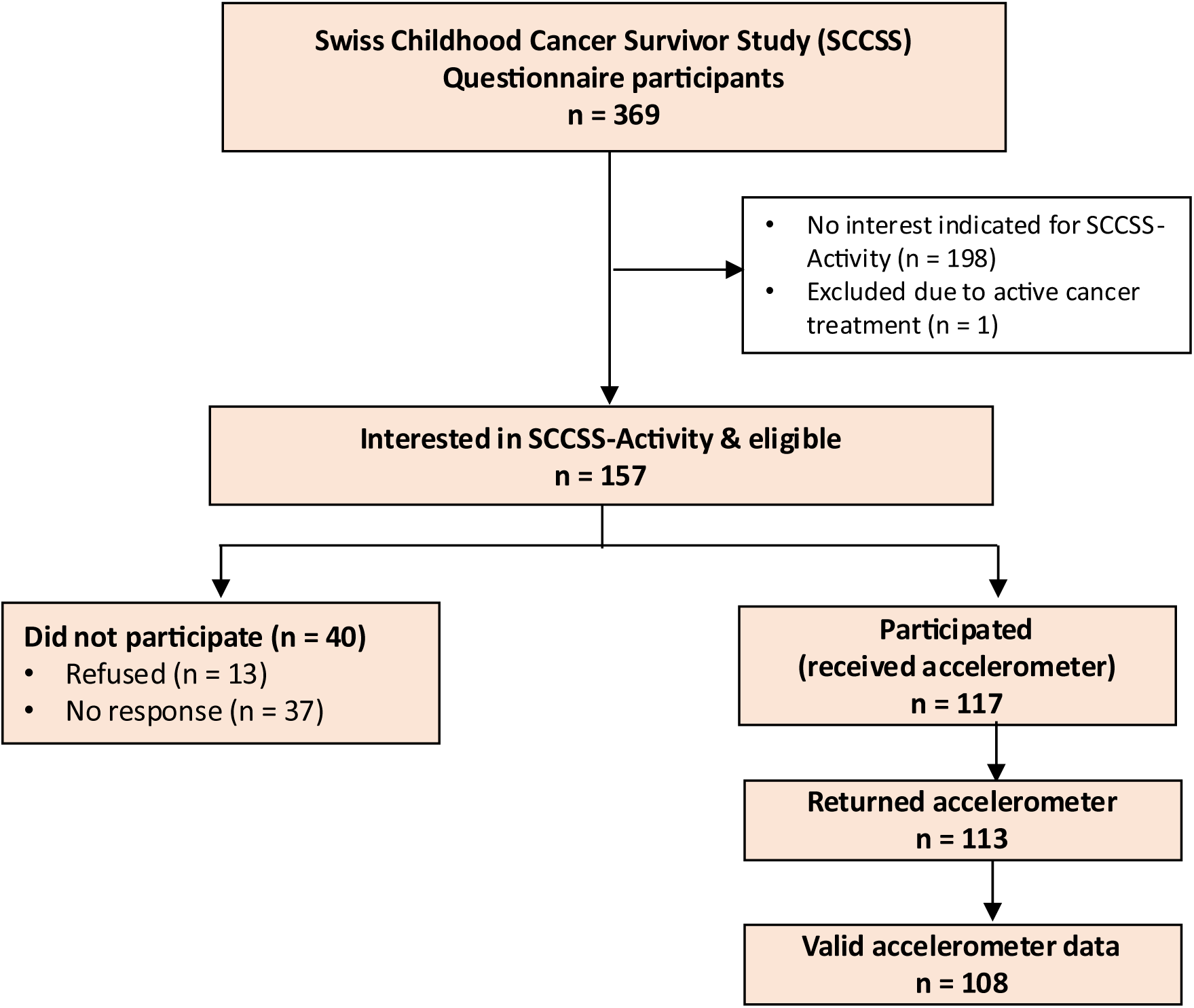
Study population tree of the SCCSS-Activity Study.

**Supplementary Table 1.** Comparison between participating and non-participating childhood cancer survivors of SCCSS-Activity.

|  | Non-participants<br>SCCSS-Activity<br>N = 261 | Participants<br>SCCSS-Activity<br>N = 108 | *p* |
| --- | --- | --- | --- |
| **Socio-demographic characteristics** |  |  |  |
| *Age at time study [median, IQR]* | 15 [11-21] | 14 [11-21] | 0.704 |
| *Age group* |  |  | 0.568 |
| 5-17 years | 156 (60%) | 68 (63%) |  |
| ≥18 years | 105 (40%) | 40 (37%) |  |
| *Female* | 121 (46%) | 54 (51%) | 0.524 |
| *Language* |  |  | 0.210 |
| German | 171 (65%) | 78 (73%) |  |
| French/Italian | 90 (35%) | 30 (28%) |  |
| *Migration background* | 53 (20%) | 18 (17%) | 0.420 |
| *Parental education\** |  |  | 0.119 |
| Primary education | 11 (4%) | 1 (1%) |  |
| Secondary education | 90 (35%) | 32 (30%) |  |
| Tertiary education | 155 (61%) | 75 (69%) |  |
| Missing | 5 (2%) | 0 (0%) |  |
| *Living location* |  |  | 0.226 |
| Rural | 51 (20%) | 15 (14%) |  |
| Town | 63 (24%) | 34 (31%) |  |
| City | 147 (56%) | 59 (55%) |  |
| **Questionnaire-reported physical activity** |  |  |  |
| Engages in exercise | 211 (81%) | 92 (85%) | 0.322 |
| Physical problem hindering exercise | 23 (9%) | 14 (13%) | 0.216 |
| Active commuting | 183 (70%) | 87 (81%) | 0.039 |
| **Cancer-related characteristics** |  |  |  |
| *ICCC3 main group* |  |  | 0.871 |
| Leukemias | 83 (32%) | 37 (34%) |  |
| Lymphomas | 42 (16%) | 14 (13%) |  |
| CNS tumors | 60 (23%) | 24 (22%) |  |
| Other | 76 (29%) | 33 (31%) |  |
| *Age at diagnosis, median [IQR]* | 8 [3-13] | 6 [3-13] | 0.620 |
| *Time since diagnosis, median [IQR]* | 7 [7-9] | 8 [7-9] | 0.753 |
| *Radiotherapy* | 47 (18%) | 26 (24%) | 0.183 |
| *Chemotherapy* | 187 (72%) | 78 (72%) | 0.911 |
| *Surgery* | 172 (66%) | 67 (62%) | 0.480 |
| *HSCT* | 14 (5%) | 8 (8%) | 0.451 |
| *Relapse* | 1 (<1%) | 0 (0%) | NA |

ICCC3 = International Classification of Childhood Cancer, 3<sup>rd</sup> edition, CNS = Central nervous system, HSCT = Hematopoietic stem cell transplantation, NA = not applicable, p-value is based upon chi-square test; \* based upon the highest degree obtained by mother or father

**Supplementary Table 2.** Time spent in physical activity intensities and sedentary behavior relative to the accelerometer wear time during waking hours (A) and relative to the total accelerometer wear time (B)

|  | Children<br>(5-17 years)<br>N = 68 | Adults<br>(≥ 18 years)<br>N = 40 | Overall<br>N = 108 |
| --- | --- | --- | --- |
| **Time spent relative to accelerometer wear time during waking hours**<br>**(A)** |  |  |  |
| **Physical activity intensities** |  |  |  |
| *Light PA without standing (%)* |  |  |  |
| Median [IQR] | 13 [11-15] | 9 [6-12] | 11 [9-14] |
| Mean (SD) | 13 (3) | 9 (3) | 11 (4) |
| *Light PA with standing (%)* |  |  |  |
| Median [IQR] | 26 [24-29] | 29 [20-34] | 26 [22-31] |
| Mean (SD) | 26 (6) | 28 (9) | 27 (7) |
| *MVPA (%)* |  |  |  |
| Median [IQR] | 10 [6-12] | 8 [7-11] | 9 [6-12] |
| Mean (SD) | 10 (4) | 9 (3) | 9 (4) |
| **Standing (%)** |  |  |  |
| Median [IQR] | 14 [12-15] | 19 [14-24] | 14 [12-18] |
| Mean (SD) | 14 (5) | 19 (7) | 16 (6) |
| **Sedentary behavior (%)** |  |  |  |
| Median [IQR] | 64 [58-69] | 60 [55-73] | 64 [57-71] |
| Mean (SD) | 64 (8) | 63 (11) | 64 (9) |
| **Time spent relative to total accelerometer wear time**<br>**(B)** |  |  |  |
| **Physical activity intensities** |  |  |  |
| *MVPA (%)* |  |  |  |
| Median [IQR] | 6 [4-7] | 6 [4-7] | 6 [4-7] |
| Mean (SD) | 6 (2) | 6 (2) | 6 (2) |
| *Light PA without standing (%)* |  |  |  |
| Median [IQR] | 8 [6-9] | 6 [4-8] | 7 [6-9] |
| Mean (SD) | 8 (2) | 6 (3) | 7 (2) |
| *Light PA with standing (%)* |  |  |  |
| Median [IQR] | 16 [15-18] | 19 [13-23] | 17 [13-20] |
| Mean (SD) | 17 (4) | 19 (7) | 17 (5) |
| **Standing (%)** |  |  |  |
| Median [IQR] | 8 [7-10] | 12 [7-16] | 9 [8-11] |
| Mean (SD) | 9 (3) | 12 (5) | 10 (4) |
| **Sedentary behavior (%)** |  |  |  |
| Median [IQR] | 40 [36-43] | 42 [36-45] | 41 [36-44] |
| Mean (SD) | 40 (6) | 41 (7) | 40 (6) |
| **Sleep (%)** |  |  |  |
| Median [IQR] | 32 [29-35] | 30 [28-34] | 31 [29-34] |
| Mean (SD) | 32 (4) | 31 (6) | 31 (5) |

**Supplementary Table 3.** Physical activity, sedentary behavior, and sleep descriptives of 6-17-year-old childhood cancer survivors (N = 68) post-stratified by age and sex to allow direct comparison with MoMo-Study [49].

|  | Weighted mean | 95%CI | Weighted median | IQR |
| --- | --- | --- | --- | --- |
| **Physical activity types** |  |  |  |  |
| *Walking (min/day)* | 91 | 85-97 | 83 | 74-106 |
| *Running (min/day)* | 9 | 7-11 | 7 | 3-14 |
| *Stair walking (min/day)* | 13 | 11-14 | 12 | 8-17 |
| *Cycling (min/day)* | 8 | 6-10 | 6 | 2-9 |
| *Moving (min/day)* | 76 | 69-82 | 74 | 58-88 |
| *Other (min/day)* | 2 | 1-3 | 1 | 0-2 |
| *All movement activities (min/day)* | 196 | 181-211 | 194 | 154-239 |
| *Steps per day* | 13,781 | 12622-14,940 | 13,543 | 10,322-16,443 |
| **Physical activity intensities** |  |  |  |  |
| *Light physical activity without standing (min/day)* | 112 | 103-121 | 109 | 93-132 |
| *Light physical activity including standing (min/day)* | 238 | 223-252 | 231 | 209-266 |
| *Moderate-to-vigorous physical activity (min/day)* | 86 | 77-95 | 88 | 54-107 |
| **Standing (min/day)** | 125 | 117-134 | 121 | 108-137 |
| **Sedentary behavior (min/day)** | 575 | 553-598 | 575 | 506-626 |
| **Sleep\* (min/day)** | 460 | 445-474 | 466 | 419-498 |

\* The MoMo study defined sleep in their publication as sleep onset to last awakening within bedtime. The authors shared the estimates for total sleep as defined in our study: mean = 439 (SD = 63), median = 443 [IQR 186-677]

**Supplementary Table 4.** Time and frequencies of sedentary bouts and frequency of sit-to-stand transitions of childhood cancer survivors.

|  | Children & adolescents<br>(5-17 years)<br>N = 68 | Adults<br>(≥ 18 years)<br>N = 40 | Overall<br>N = 108 |
| --- | --- | --- | --- |
| *Time spent on sedentary bouts ≥30min*<br>*(min/day)* |  |  |  |
| Median [IQR] | 226 [159-320] | 319 [230-416] | 264 [172-350] |
| Mean (SD) | 242 (109) | 325 (117) | 273 (118) |
| *Frequency (count) of sedentary bouts >30 min*<br>*(N/day)* |  |  |  |
| Median [IQR] | 5 [3-6] | 5 [4-7] | 5 [4-6] |
| Mean (SD) | 5 (2) | 6 (2) | 5 (2) |

**Supplementary Table 5.** Proportion of survivors fulfilling guidelines or recommendations for physical activity, steps, sleep or sedentary behavior.

|  | Children<br>(5-17 years)<br>N = 68 |  | Adults<br>(≥ 18 years)<br>N = 40 |  | Overall<br>N = 108 |  |
| --- | --- | --- | --- | --- | --- | --- |
|  | N (%) | 95%CI | N (%) | 95%CI | N (%) | 95%CI |
| *Physical activity guidelines* | 49 (72%) | 60-82 | 39 (98%) | 83-100 | 88 (82%) | 73-88 |
| *Sleep guidelines* | 29 (43%) | 31-55 | 26 (65%) | 48-79 | 55 (51%) | 41-60 |
| *Sedentary behavior recommendations* | 7 (10%) | 5-20 | 11 (28%) | 16-44 | 18 (17%) | 11-25 |
| *Combined guideline adherence* | 6 (9%) | 4-19 | 8 (20%) | 10-36 | 14 (13%) | 8-21 |

Physical activity guideline adherence: ≥60 minutes MVPA/day for children; ≥150 minutes MVPA/week for adults<sup>4</sup>
Sleep guidelines adherence: ≥9 hours/night for 6-13 years year-olds, ≥8 hours/night for 14-17 year-olds; ≥7 hours/night for adults.<sup>41</sup>
Sedentary behavior guideline adherence: <8 hours/day sedentary for children, <9 hours/day sedentary for adults<sup>4,44</sup>
Combined guideline adherence: Adherence to physical activity, sleep, and sedentary behavior guidelines simultaneously

